# Double Mammary In Situ: Predicting Feasibility of Right Mammary Artery In Situ for Circumflex Coronary Artery System

**DOI:** 10.1101/2020.12.05.20243964

**Authors:** Rafik Margaryan, Daniele Della Latta, Giacomo Bianchi, Nicola Martini, Andrea Gori, Marco Solinas

**Author notes:** Correspondence concerning this article should be addressed to Rafik Margaryan, MD, PhD, Via Aurelia Sud 303.

## Abstract

**Objectives:** Double (bilateral) mammary artery in situ revascularization seems less attractive to surgeons because of limited mammary length of right mammary artery, scare, or no means of its length estimation.

**Methods:** We’ve selected patients who have used bilateral mammary artery for revascularization and divvied them into two groups: in situ and y-graft groups. We have used preoperative chest x-rays to build a predictive model with neural networks that could predict feasibility of in situ bilateral mammary revascularization.

**Results:** The predictive model was able to predict a positive outcome with 96% percent accuracy (p < 0.01). Models sensitivity and specificity were 96% and 95% respectively. Neural networks can be used to predict double mammary feasibility using chest x-rays. Model is capable of predicting positive outcomes with 95% accuracy.

**Conclusions:** Chest x-ray base model can accurately predict the feasibility of in situ bilateral mammary artery revascularization.

## Introduction

The use of double (bilateral) internal mammary arteries (BIMA) is scarcely diffused in the real world (ElBardissi et al., 2012; LaPar et al., 2015), despite being supported by robust evidence over the last two decades. Right internal mammary artery (RIMA) is anatomically, histologically and functionally similar to left mammary artery (LIMA), also in terms of endothelial function and RNA expression level (Ferrari et al., 2014; Märkl, Raab, Arnholdt, & Vicol, 2003).

Lack of standardization for the appropriate use of the right internal mammary artery (RIMA) has limited its routine use in myocardial revascularization. RIMA can be anastomosed in situ to the left anterior descending artery (LAD) or to the proximal branches of the circumflex artery, but its length may pose technical challenges. Although skeletonization is a technique that provides better internal mammary artery (IMA) length, it is not widely accepted. As a result, it is not always possible to choose the best anastomotic site, and we are forced to graft the RIMA according to its length rather than the most suitable coronary artery segment. Alternatively, the RIMA can be used as a free graft, which is anastomosed to the left internal mammary artery (LIMA) as a composite conduit. This option is more technically demanding and can expose to higher rates of competitive flow with increased graft failure (Pevni et al., 2007) though no definitive data are available to confirm this finding (Glineur et al., 2008). Mid-term clinical outcome seems to be similar regardless of BIMA configuration, either in situ or as a composite conduit (Glineur et al., 2008). In situ double mammary grafting is safe and durable bypass strategy when circumference artery system has suitable targets, although personal judgment of right mammary length, hence, RIMA in situ feasibility remains subjective.

We sought do develop prediction model using chest x-ray and deep neural networks in orders to estimate feasibility of right mammary artery to circumflex artery targets (early obtuse marginal branches and intermediate/early diagonal arteries)

## Material and Methods

### Patients

We have selected patients from our electronic health record system, undergoing double mammary artery grafting from 2007 until august 2019 (n = 292) were selected for this study. All patients underwent successful on-pump bypass surgery. For this analysis we’ve ignored other types of bypasses. We divided the population into two groups:

- In situ bypass grafting
- Grafting using y- or t-grafts

127 were double mammary grafts in-situ and 165 were double mammary grafts Y grafts. Mean age of the patients was 61.4 *±* 7.7. All other demographic data are presented in Table 1. All patients had preoperative chest x-ray (antero-posterior and lateral).

**Table 1.** Patient Characteristics

| **\*\*Characteristic\*\*** | **\*\*In Situ\*\*, N = 127** | **\*\*Y-graft\*\*, N = 165** | **\*\*p-value\*\*** |
| --- | --- | --- | --- |
| Patient Age | 61 (8) | 62 (8) | 0.8 |
| BMI | 27.1 (4.1) | 27.0 (5.0) | 0.8 |
| Hypertension | 81 / 127 (64%) | 100 / 165 (61%) | 0.6 |
| Hypercholesterolemia | 73 / 127 (57%) | 95 / 165 (58%) | >0.9 |
| Diabetes | 24 / 127 (19%) | 40 / 165 (24%) | 0.3 |
| Diabetes on Insulin | 1 / 127 (0.8%) | 4 / 165 (2.4%) | 0.3 |
| Sex |  |  | 0.7 |
| Female | 8 / 127 (6.3%) | 12 / 165 (7.3%) |  |
| Male | 119 / 127 (94%) | 153 / 165 (93%) |  |
| Preoperative Creatinine | 0.93 (0.21) | 0.95 (0.52) | 0.7 |
| ON Pump | 112 / 127 (88%) | 140 / 165 (85%) | 0.4 |
| CPB Time | 101 (29) | 100 (34) | >0.9 |
| XC Time | 70 (22) | 70 (25) | 0.9 |

### Data Acquisition and Processing

292 chest digital X-ray examinations (DX) were performed on patients enrolled for valvular surgery. X-ray images of lateral latero (LL) projections were used to train a deep convolutional neural network (DNN). All DX were taken with standing patient placed 180 cm from the X-ray tube using a digital flat detector panel with a spatial resolution of 0.139×0.139 mm^2^. The size of the image matrix is variable (2500×2500-4000×4000 px) depending on the size of the patient’s chest as well as the X-ray exposure. The potential difference (kV) and exposure current (mAs) parameters are set automatically of the X-ray scanner in order to obtain an adequate contrast independently of the variation of the attenuation of the body of each patient. The dataset was splitted into 183 images to train the DNN and 22 to test the learning. The size of all the images was readjusted to 2000×2000 with a centered crop operation so as to maintain the DNN input layer with fixed size and partially mask the background that does not provide an useful information for the classification task. Finally, to minimize the computational load for network training, the images were scaled to 512×512 px. Whole data was divided randomly into two parts: train dataset and test dataset. Train dataset was used to create the model and validate it. Test dataset was used for only external validation and never has been used for tuning or improving the algorithm.

### Network Architecture

A GoogLeNet network (Szegedy et al., 2014) was used to perform the classification task. The outline of the proposed architecture was reported in Table 1. The network comprises 22 trainable layers and is made by nine Inception module stacked upon each other. A single Inception module represents a combination of 1×1, 3×3 and 5×5 convolutional kernels, used in parallel with the aim to progressively cover a bigger area and extract details (1×1) as more general features (5×5). The results are then concatenated with the outcome of a max-pooling layer and passed to the next layer. The presence of 1×1 convolutions before the 3×3, 5×5 convolutions and after the pooling layer help to reduce the number of parameters in the Inception module and add more non-linearities to the network. To apply the GoogLeNet network architecture to the surgical task some implementation changes had to be made: 1) the input layer dimensions, set at 512 × 512, with a number of channels equals to one instead of three, being the inputs grayscale images; 2) in the output layer only two units were used, as we had to classify the images according the two possible locations of surgical incision; The network was designed and trained with TensorFlow (Workshop on Wireless Traffic Measurements and Modeling, USENIX Association, ACM SIGMOBILE, ACM Special Interest Group in Operating Systems, & ACM Digital Library, 2016), running for 500 epochs on a single NVIDIA Titan Xp. Stochastic gradient descent and Adam optimizer were used to minimize a categorical cross-entropy for the network parameters update.

Schematic representation of the GoogLeNet network used to classify the input images. For a detailed description of the network architecture see (Szegedy et al., 2014).

The size of all the images was readjusted to 2000×2000 with a centered crop operation so as to maintain the DNN input layer with fixed size and partially mask the background that does not provide an useful information for the classification task. Finally, to minimize the computational load for network training, the images were scaled to 512×512 px. To evaluate the progress of learning the accuracy was calculated between the predicted and the real “point of incision.”

## Results

There were no hospital or 30 days mortality. In original model antero-posterior and lateral chest x-rays were include. However, lateral images were not changing or changing little of predictive value. The final model has only antero-posterior chest x-rays. Model has predicted with 95.55% accuracy (p < 0.01). There were no significant differences in comorbidities divided by type of graft and sex group(see Table 1. Model’s sensitivity and specificity were 0.96 and 0.95. Area under curve (AUC) was 0.96. Predicted type and real type of configuration is reported in the Table 2. All combinations of grafts and type of revascularization see Table 3.

**Table 2.** True and Predicted Confituration

| **\*\*Characteristic\*\*** | **\*\*In Situ\*\*, N = 127** | **\*\*Y-graft\*\*, N = 165** | **\*\*p-value\*\*** |
| --- | --- | --- | --- |
| Predicted |  |  | <0.001 |
| In Situ | 122 / 127 (96%) | 8 / 165 (4.8%) |  |
| Y-graft | 5 / 127 (3.9%) | 157 / 165 (95%) |  |
| Sex |  |  | 0.7 |
| Female | 8 / 127 (6.3%) | 12 / 165 (7.3%) |  |
| Male | 119 / 127 (94%) | 153 / 165 (93%) |  |

**Table 3.** Graft Configuration vs. Anatomical Vassel Distribution

| **Characteristic** | **In Situ**, N = 127 | **Y-graft**, N = 165 | **p-value** |
| --- | --- | --- | --- |
| Left Anterior Descending | 126 / 127 (99%) | 165 / 165 (100%) | 0.3 |
| Intermedial Branch | 47 / 127 (37%) | 32 / 165 (19%) | <0.001 |
| First Marginal | 46 / 127 (36%) | 31 / 165 (19%) | <0.001 |
| Second Marginal | 7 / 127 (5.5%) | 1 / 165 (0.6%) | 0.011 |
| Sex |  |  | 0.7 |
| Female | 8 / 127 (6.3%) | 12 / 165 (7.3%) |  |
| Male | 119 / 127 (94%) | 153 / 165 (93%) |  |

## Discussion

In this study we were able to build a predictive model using chest x-rays which does not require any type of measurement other then raw chest x-ray data with very high performs. To our knowledge it is first of its kind and it seeks to resolve an actual clinical problem. Absolute length of the bilateral mammary arteries is an intrinsic feature, and as we have demonstrated can be extracted if necessary. Hence, using medical imaging data is most accurate way to estimate intrinsic length of mammary arteries. Deep Learning models in general have very high good performance, and in this clinical scenario it has very high accuracy and AUC values for predicting possible double mammary in-situ bypass grafting. It proves to have hit sensitivity and specificity. It is particularly valuable for total arterial revascularization. Certainly it is bias by its nature of being single center. Future commitment for international chest x-ray database should be created for better model.

Bilateral independent mammary arteries is most efficient and durable solution as state of the art in complete atrial revascularization(Head, Milojevic, Taggart, & Puskas, 2017). There are reports in literature that configuration does not change the mortality (Di Mauro et al., 2016). In any case when one does not want to comprise the left mammary artery and/or has a doubt on flows and distal resistances, the independent in situ configuration is the right choice to our experience and knowledge.

## Conclusion

Neural networks can be used to predict double mammary feasibility using chest x-rays. Model is capable of predicting positive outcome with 95.55% accuracy. It is unique, non operator dependent method which can be very valuable in clinical situations.

## Data Availability

All data will be available after deidentification.

---

**Figure 1.**
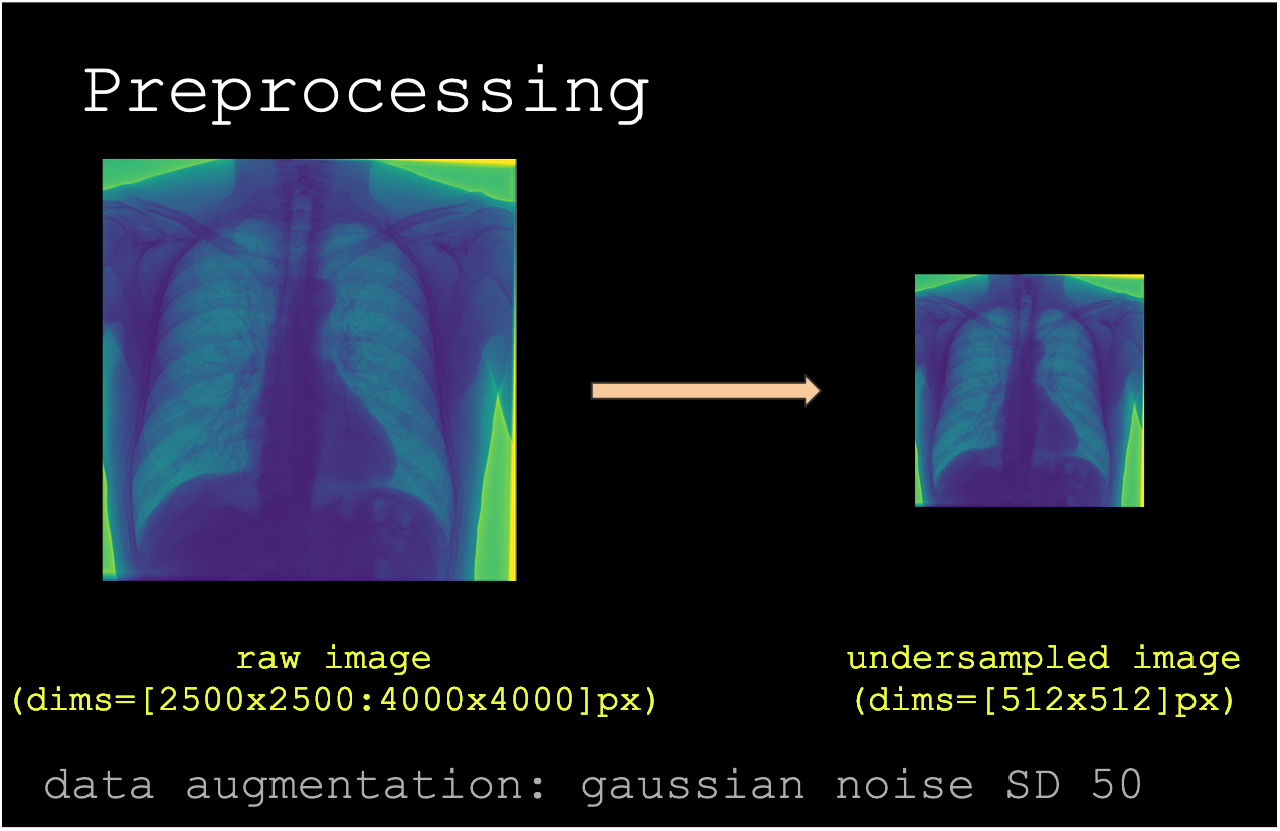
Figure preprocessing before feeding into neural networks.

**Figure 2.**
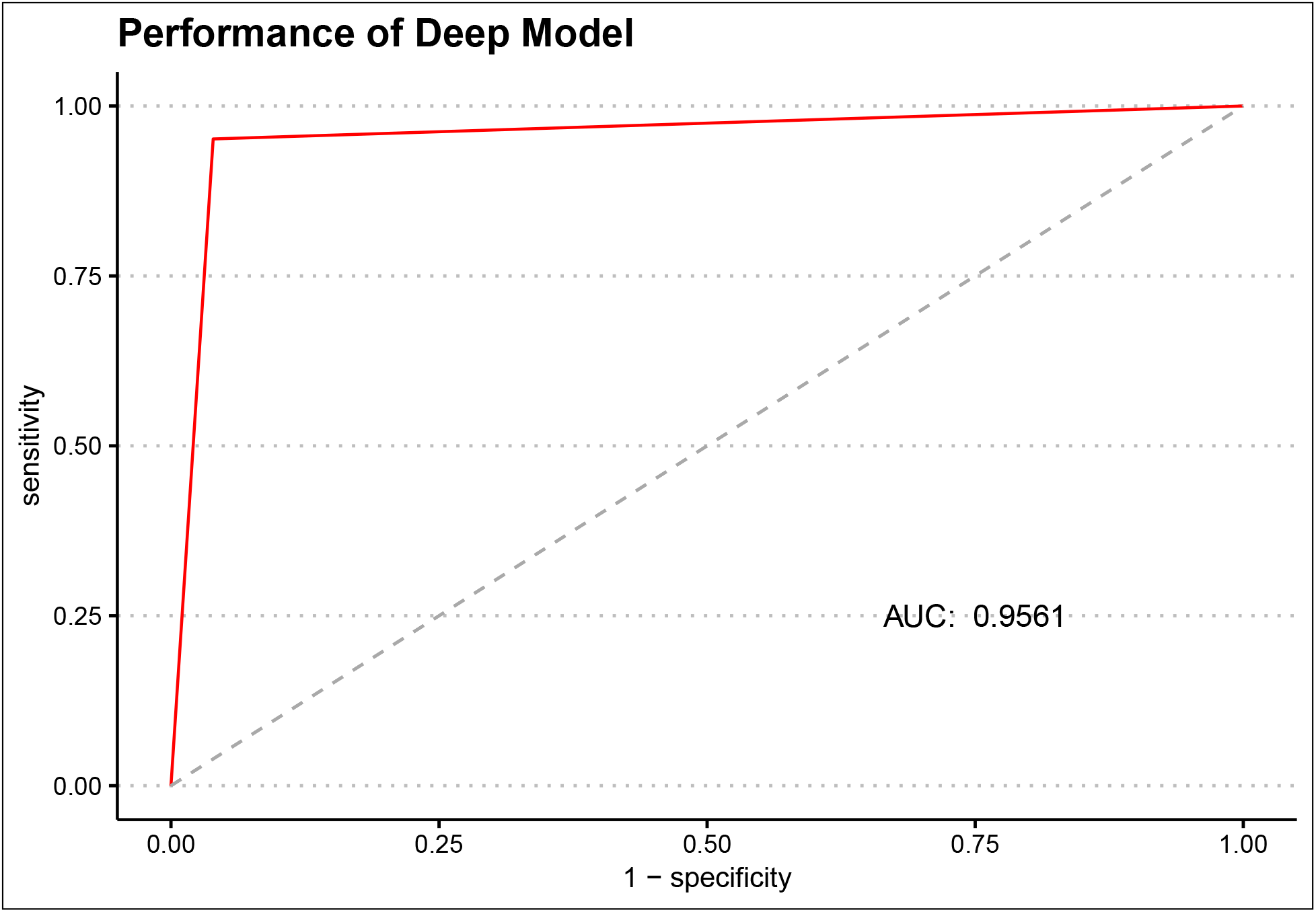
Model performance expressed with ROC (receiver operating characteristic) curve.

